## Supplementary File 2-sFigure for "Genetic associations between circulating immune cells and periodontitis highlight the prospect of systemic immunoregulation in periodontal care"

**Supplementary Figures**

**Figure of Contents**

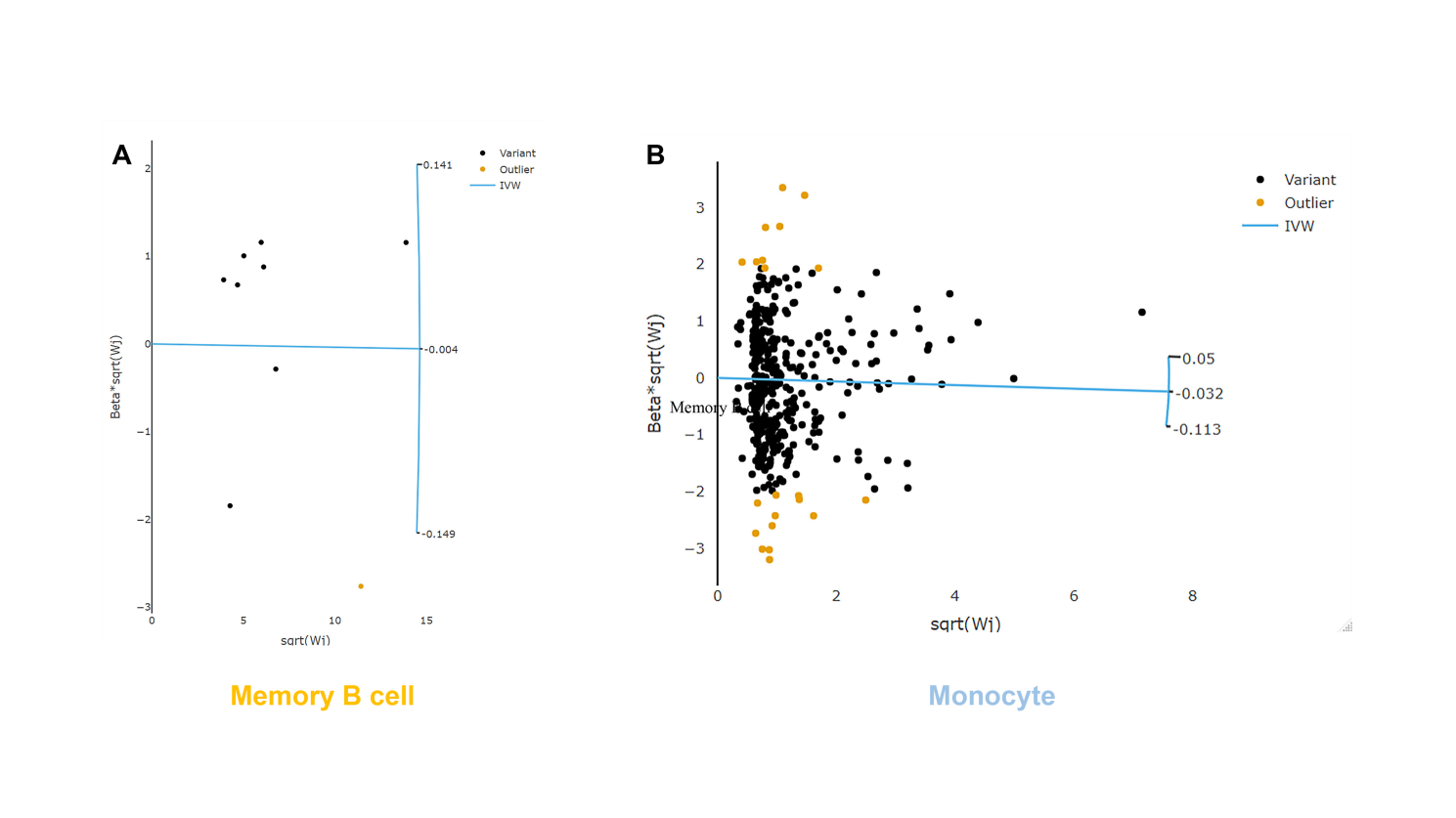

**Figure s1****.** Scatter plot to explore outliers for two features with considerable heterogeneity. Based on the RadialMR method, we detected 1 and 21 outlier SNPs from the memory B cell **(A)** and the monocyte **(B)**, respectively.

**Abbreviations:** IVW, inverse variance weighted; SNP, single nucleotide polymorphism.

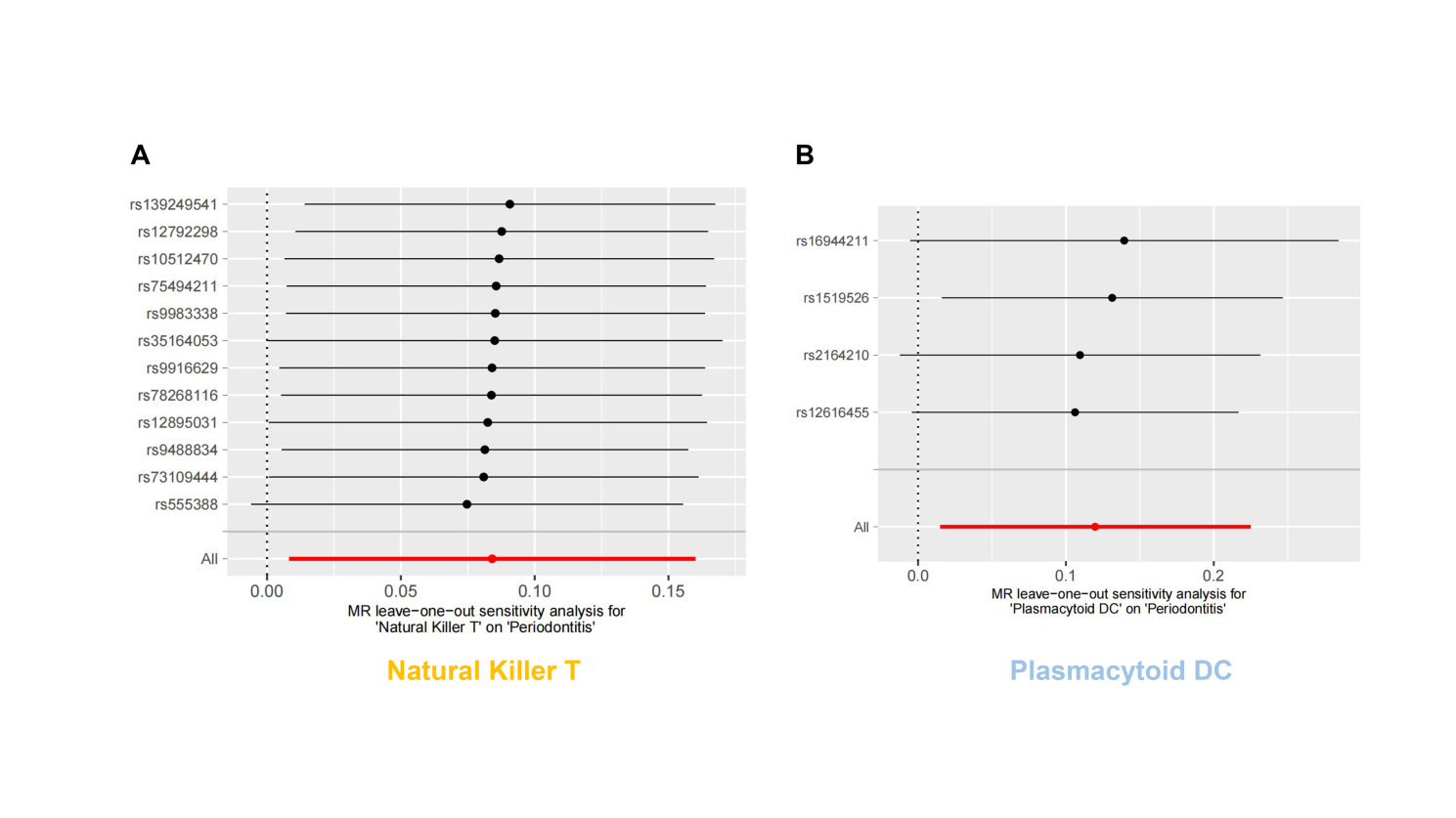

**Figure s2.** Results of leave-one-out sensitivity analysis. No influential SNPs were detected for either the Natural Killer T cell **(A)** or the plasmacytoid DC **(B)**.

**Abbreviations:** DC, dendritic cell; MR, Mendelian randomization; SNP, single nucleotide polymorphism.

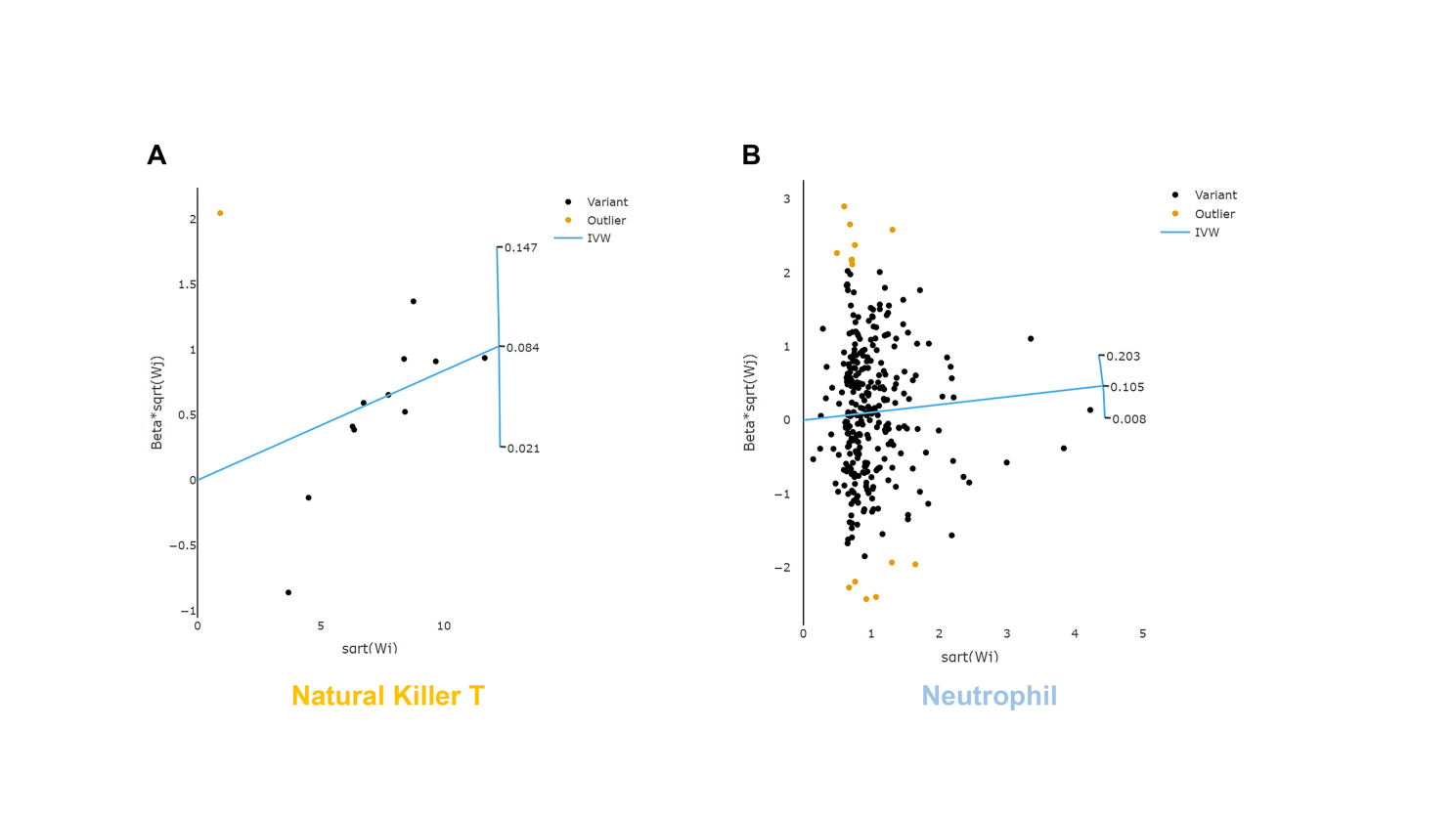

**Figure s3.** Scatter plot to explore outliers for significant results. Based on the RadialMR method, we detected 1 and 14 outlier SNPs from the Natural Killer T cell **(A)** and the neutrophil **(B)**, respectively.

**Abbreviations:** IVW, inverse variance weighted; MR, Mendelian randomization; SNP, single nucleotide polymorphism.

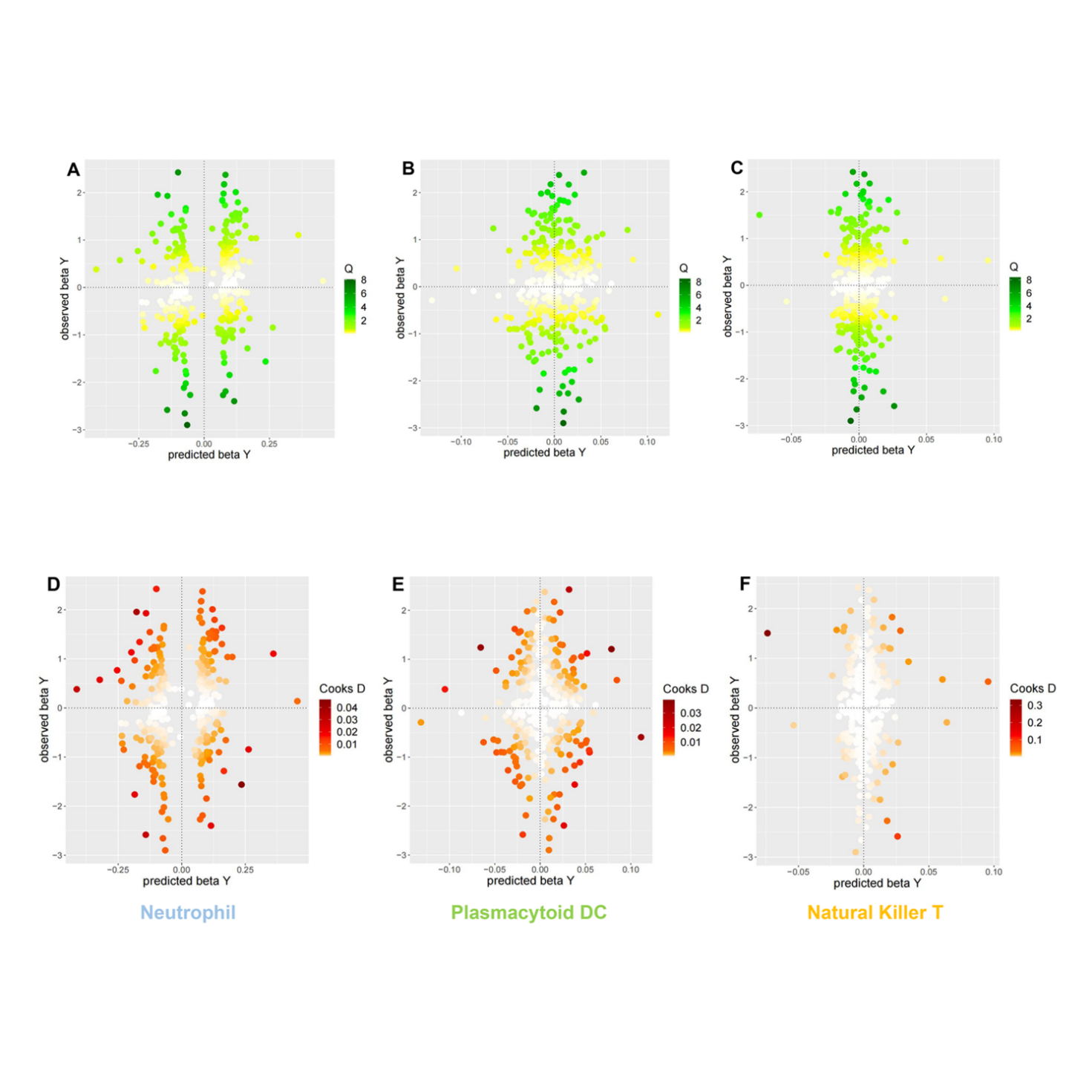

**Figure s4.** Scatter plot of the Cochran's Q test and Cook's distance to explore outlier or influential variations in MR-BMA. Panels **(A-C)** illustrate the application of Cochran's Q test in evaluating the effect of neutrophils, plasmacytoid DCs, and Natural Killer T cells on periodontitis, respectively. Panels **(D-F)** illustrate the application of Cook's distance in evaluating the effect of neutrophils, plasmacytoid DCs, and Natural Killer T cells on periodontitis, respectively.

**Abbreviations:** BMA, Bayesian model averaging; DC, dendritic cell.

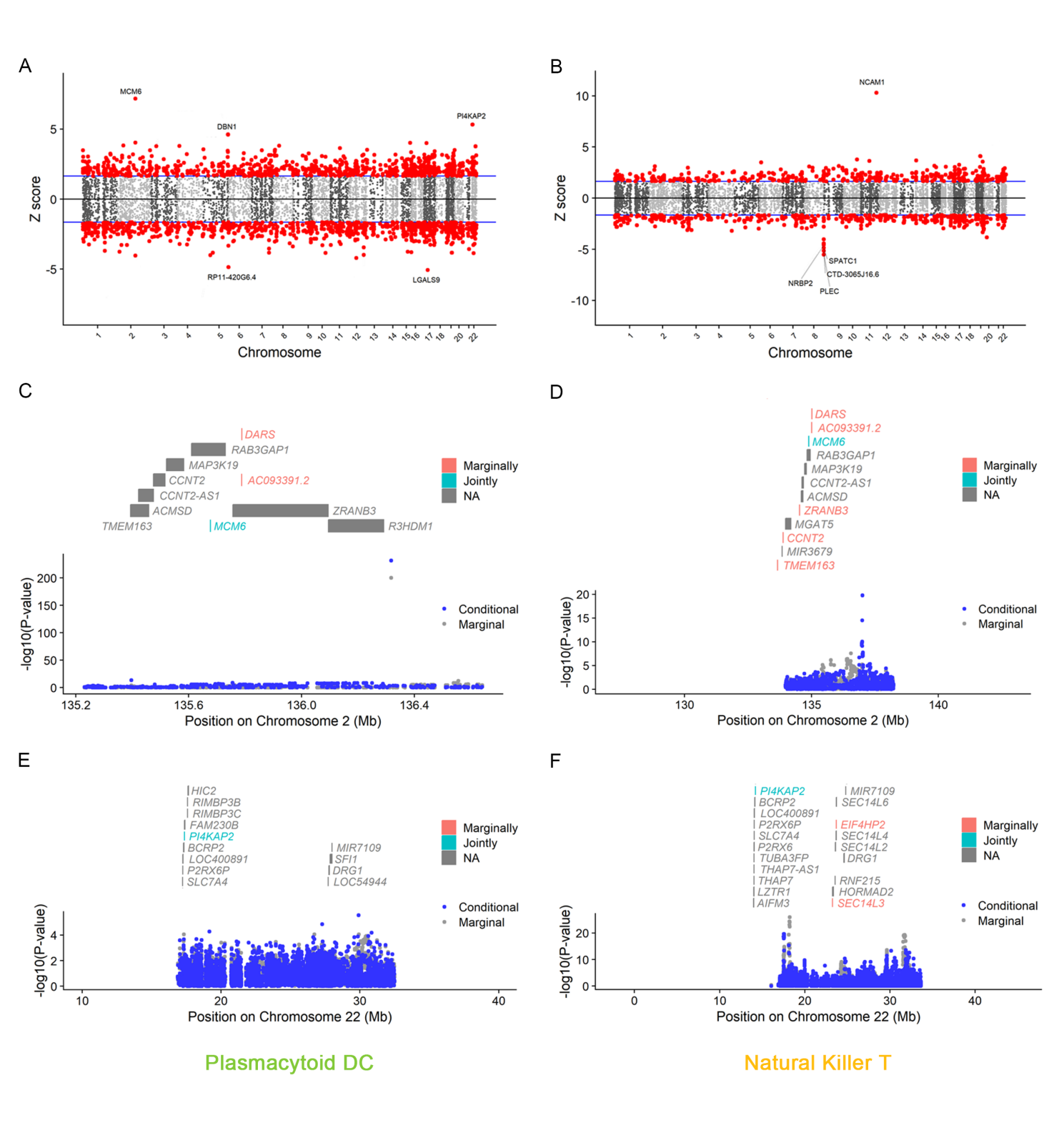

**Figure s5.** Results of the TWAS and colocalization analysis. **(A-B)** The Manhattan plot illustrates gene-trait associations for plasmacytoid DC **(A)**, and Natural Killer T cell **(B)**. The x-axis represents the genomic position. The blue lines indicate a Z-score of 1.96. The red circles denote significant gene-trait associations (*P* < 0.05). Ten genes satisfy a multiple corrected threshold of *P* < 6.27×10^-6^. **(C-F)** The regional Manhattan plot demonstrates the conditional analysis for *MCM6* in plasmacytoid DC **(C)**, Natural Killer T cell **(D)**; as well as *P14KAP2* in plasmacytoid DC **(E)**, Natural Killer T cell **(F)**. The grey bars mark the location of genes on the chromosome. The genes highlighted in orange and green on the graph represent the marginally and jointly significant genes that best explain the GWAS signals. The gray and blue dots represent GWAS *p*-value before and after conditioning on the jointly significant gene.

**Abbreviations:** DC, dendritic cell; GWAS, genome-wide association study; PP, posterior probability; TWAS, transcriptome-wide association study.
